# Flooding and malaria in the Sahara: description of the 2024 outbreak in the Kidal region, Mali

**DOI:** 10.1101/2025.08.14.25333744

**Authors:** AM Dolo, M Cissoko, A Teme, K Keita, M Sanogo, CAT Traoré, M Magassa, A Koné, FA Roy, I Sagara, J Gaudart

**Affiliations:** Malaria Research and Training Center (MRTC), FMOS & FAPH, USTTB, BP 1805 Bamako, Mali; Aix Marseille University (AMU), French National Institute of Health and Medical Research (INSERM), Development Research Institute (IRD), 13005 Marseille, France; National Malaria Control Program (NMCP), 233, Bamako, Mali; Directorate General of Health and Public Hygiene (DGHPH), Bamako BP 233, Bamako, Mali; Kidal Regional Health Direction, Mali; Univ. Boni, Digital health & innovation research unit, LaMIC-SD, Bobo Dioulasso, Burkina Faso; Aix Marseille University, APHM, INSERM, IRD, SESSTIM, Hop Timone, BioSTIC, Biostatistic & TIC, 13005 Marseille, France

**Keywords:** outbreak, Sahara, malaria, climate change, extreme events, floods, Mali

## Abstract

**Context:** During the 2024 rainy season, extreme rainfall events caused flooding in Mali, leading to an outbreak of malaria in the Kidal region of northern Mali. The aim of this study was to describe this epidemic and the response implemented by the national and regional authorities.

**Methods:** Weekly rainfall data (0.25° resolution) from TRMM and malaria cases from national health system. Data on SMCs and mobile clinics come from the national malaria control program. Malaria incidence and rainfall trends were analyzed, with the epidemic described according to severity and age group. The effects of the intervention were assessed using interrupted time series with a generalized additive model.

**Results:** Rainfall began in week 25 (June) and lasted 15 weeks, averaging 3.86 mm/day (CI95: 3.23–4.57). The malaria epidemic began in week 30, peaked at 1,014 cases in week 39, and lasted 28 weeks (incidence: 1.688/1,000 person-weeks; CI95: 1.687–1.690). The [0-11] months and [1-4] years age groups were the most affected, with respective incidences of 1.48 and 0.97 per 1000 person-weeks for severe cases, and 5.04 and 2.35 per 1000 person-weeks for uncomplicated cases. Two interventions were implemented: i) 2 SMC campaigns among children aged 0 to 4 years for the first round, and 0 to 10 years for the second round, during weeks 31 (July) and 41 (October) respectively (6,236 and 27,334 children respectively); ii) mobile teams deployed from week 34 (August) to 39 (September) in remote areas and Internally Displaced People (IDP) sites, with 35 members in addition to the 36 community health workers. These interventions significantly reduced morbidity, with respective SIRs of 0.5 CI95[0.33; 0.75] (p=0.002) and 0.48 CI95[0.28; 0.82] for SMC and mobile teams.

**Conclusion:** Climate change is increasing extreme weather events like floods, altering malaria risk in unstable areas such as northern Mali. Preparedness should include reinforcing community health workers, ensuring adequate diagnostic and treatment stocks, and deploying targeted mobile teams.

## 1. Introduction

The regions of northern Mali, south of the Sahara, present an environmental context conducive to unstable malaria transmission. Malaria occurs in the form of epidemic outbreaks when rainfall exceeds the usual average (1), with a high number of serious cases and deaths. According to the World Health Organisation (WHO) report, extreme weather events linked to climate change, such as flooding, which should increasing the number of malaria cases (2). According to the Intergovernmental Panel on Climate Change (IPCC), the Sahel is one of the region vulnerable to the effects of climate change, because of an increase of extreme events such as extreme rainfall and temperatures rises at least 2°C in the short term (2021-2040), a rate 1.5 times higher than the global average (3). An increase in extreme rainfall in the Sahel- Saharan zone will lead to rain and river flooding (3). In the Sahara, compared with other regions of Africa, forecasts of the frequency of climatic phenomena and the onset of epidemics are still poorly known. Mali suffered 15 floods between 1980 and 2024 (4). The last year of flooding in the region, in 2024, caused extensive damage in all regions. According to the national Crisis Management and Coordination Centre, the country recorded 729 cases of flooding, 47,306 cases of collapses, the destruction of 195,845 hectares of agricultural land, and the loss of 756,127 tonnes of cereals. There were also 465,226 people affected, 33 health centres affected and 66,032 houses damaged (5). This situation led to thousands of people being displaced, exposing them to health risks. In response to this situation, the government of Mali declared a state of national disaster (decree no. 2024-0485) (6).

Extreme rainfall during the first two weeks of August 2024 caused severe flooding in the Kidal region (7). Exposing the population to an increased risk of malaria in a context of prolonged insecurity and internally displaced persons, the situation has been exacerbated, especially as 83% of the population of Kidal has no access to a formal health centre (8). These circumstances not only complicated relief efforts but also confirmed the fragility of the region in the face of malaria epidemics.

The aim of this study was to describe the outbreak of malaria during the 2024 floods in the Kidal region, Mali, and the response implemented by the national and regional authorities.

## 2. Methods

### 2.1. Study location

The study was carried out in the Kidal region of northern Mali, comprising 4 health districts (HDs). The population of Kidal in 2024 was 106,775 (RGPH-2019) (9), spread over an area of 151,430 km^2^ and around 250 km from the Algerian border and 1,674 km from the capital Bamako, in a zone of active conflict. Located in the desert eco-climatic zone, the climate is characterized by wide variations in temperature: the average temperature is around 33°C (with a minimum of 21°C and a maximum of 45°C). Average annual rainfall is 94 mm over 11 weeks (10), from June to September, and 135.11 mm over 15 weeks for 2024 (10), with 40.84 mm in 14 days, causing flooding in early August.

### 2.2. Ethical approval

Authorization to conduct this study was obtained from the Ministry of Health and Social Affairs of Mali through the National Malaria Control Program (No. 108/MSDS-SG/PNLP) on 5 February 2025, in accordance with Malian regulations on ethics and medical research, as well as approval from the Ethics Committee of Aix-Marseille University (File Ref. No: 2025-03-13- 01/DPO 760100) on 13 March 2025. A certificate of exemption from the legislation on the protection of personal data was also issued to us by the General Data Protection Regulation (GDPR) team of Aix-Marseille University.

The authors did not have access to any information that could identify individual participants during or after the data collection process.

### 2.3. Data collection and sources

#### 2.3.1. Malaria cases and population

Cases of uncomplicated and severe malaria confirmed by Rapid Diagnostic Test (RDT) and by age group (0-11 months, 1-4 years, 5-14 years, >14 years) in health facilities from 23 May 2024 to 02 February 2025 were reported to the national health information system (DHIS2). The study focused on data aggregated at the regional level and by week. The population was extracted from RGPH 2019 updated.

#### 2.3.2. Intervention data

Intervention data from SMC and mobile clinics were obtained from the National Malaria Control Department.

#### 2.3.3. Meteorological data

From May 23, 2024, to February 2, 2025, Precipitation data (mm) were extracted from Tropical Rainfall Measuring Mission (TRMM) Rainfall Estimate L3 0.25 degree x 0.25 degree V7 (TRMM_3B42) produced at the NASA GES DISC (10) on a daily basis and then cumulated on a weekly basis.

### 2.4. Data analysis

#### 2.4.1. Descriptive analysis

The incidence rate per 1000 person-week was calculated for the whole period, for all confirmed malaria cases according to severity and age group (0-11 months, 1-4 years, 5-14 years, >14 years). (11). The cumulative rainfall per week was calculated.

#### 2.4.2. Spatial analysis

None of the data reported was geolocalized. The population at risk of malaria was estimated using WorldPop. This approach enabled us to map an estimate of the spatial distribution of the population at risk of malaria.

#### 2.4.3. Regression modal

An interrupted time series analysis was implemented to study the impact of interventions on the number of malaria cases (13) using a mixed generalized additive model. A quasi-Poisson distribution was used to account for overdispersion in the number of malaria cases; time (week) was modeled by a random effect; an offset(log(pop)) was used to estimate standardized incidence ratios; the model was adjusted for rainfall with a 5-month lag (13). Non-linear relationships were modelled using spline functions. Two binary intervention variables were used, one for SMC (Seasonal Malaria Chemoprevention), considering a 28-day protective efficacy and the other for intervention mobile teams. For each intervention, the time after the start of the intervention was also modelled. The complete model, including interventions, temporality and rainfall, was used to simulate the time series of the number of cases and check consistency with observations.

Cases = offset(log(population)) + SMC + f(Time from SMC) + Mobile Clinic + f(Time from Mobile Clinic) + f(5 weeks lagged Rainfall) + random effect(Time), model = Quasi-Poisson

#### 2.4.4. Analysis software

Statistical analysis was carried out using R software (version 4.1.3). The map was produced using QGIS software (version 3.22.8).

## 3. Result

### 3.1. General description of the malaria incidence time series

A total of 6311 malaria cases were confirmed, including 2310 severe cases from week 23 of 2024 (May) to week 5 of 2025 (February). The rains began in week 25 (June) and lasted 15 weeks, with an average of 3.86 mm/day IC95[3.23; 4.57]. The malaria epidemic began around week 30 (July 2024) with 59 malaria cases, and lasted 28 weeks, peaking in week 39 (September 2024) with 1,014 cases. The incidence rate was 1.688 /1000 persons.week CI95 [1.687; 1.690].

**Figure 1:**
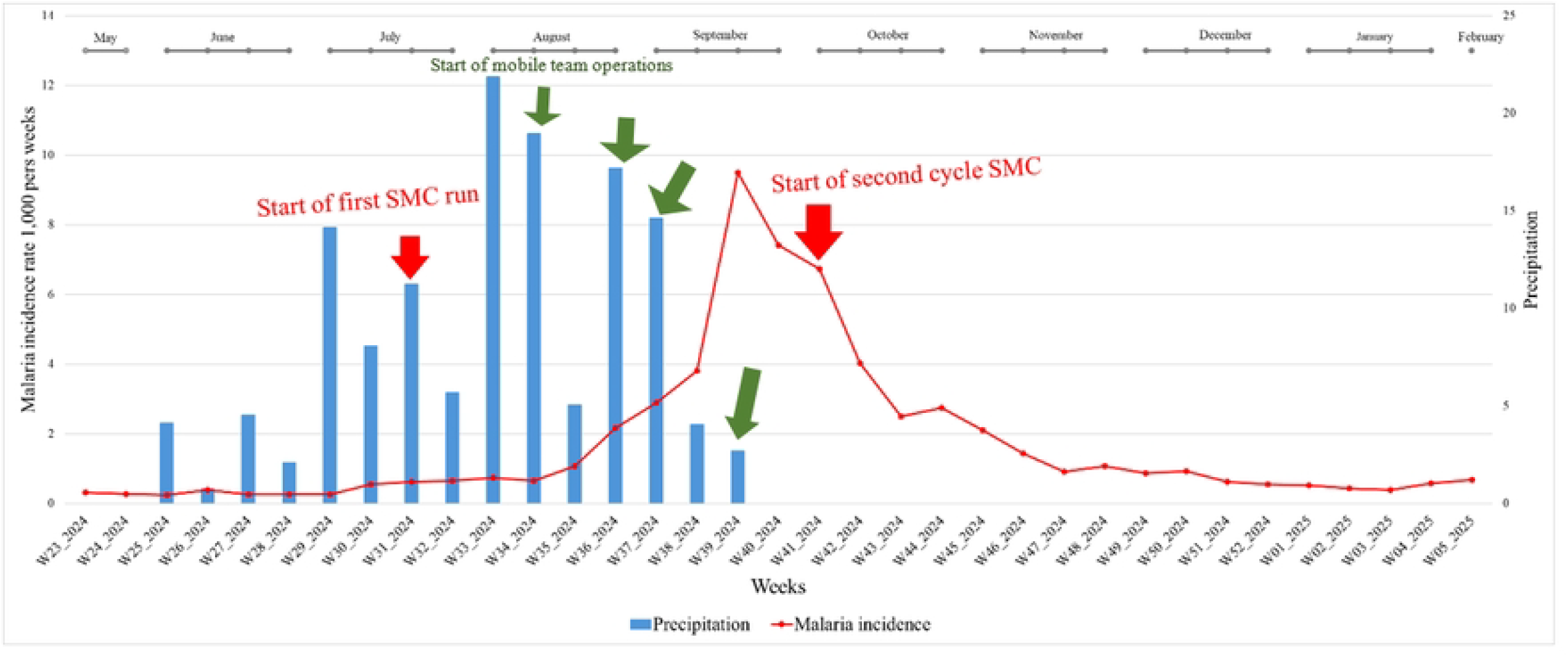
Evolution of weekly malaria incidence rate and rainfall from S23 (May-2024) to S5 (February-2025).

The red curve represents the malaria incidence rate per 1,000 person.weeks. The epidemic peak was observed in September, precisely at week 39, corresponding to the end of the rainy season. Weekly cumulative rainfall (mm) is shown in the blue bar chart.

### 3.2. General description of the time series of incidence of uncomplicated malaria by age group

A total of 4001 confirmed cases of uncomplicated malaria were recorded. The age groups most affected were children aged [0-11] months and [1-4] years, with an incidence rate of 5.04 and 2.35 per 1,000 person-weeks respectively. On the other hand, the age group least affected was [5-14] years, with an incidence rate of 0.62 per 1,000 person-weeks.

**Figure 2:**
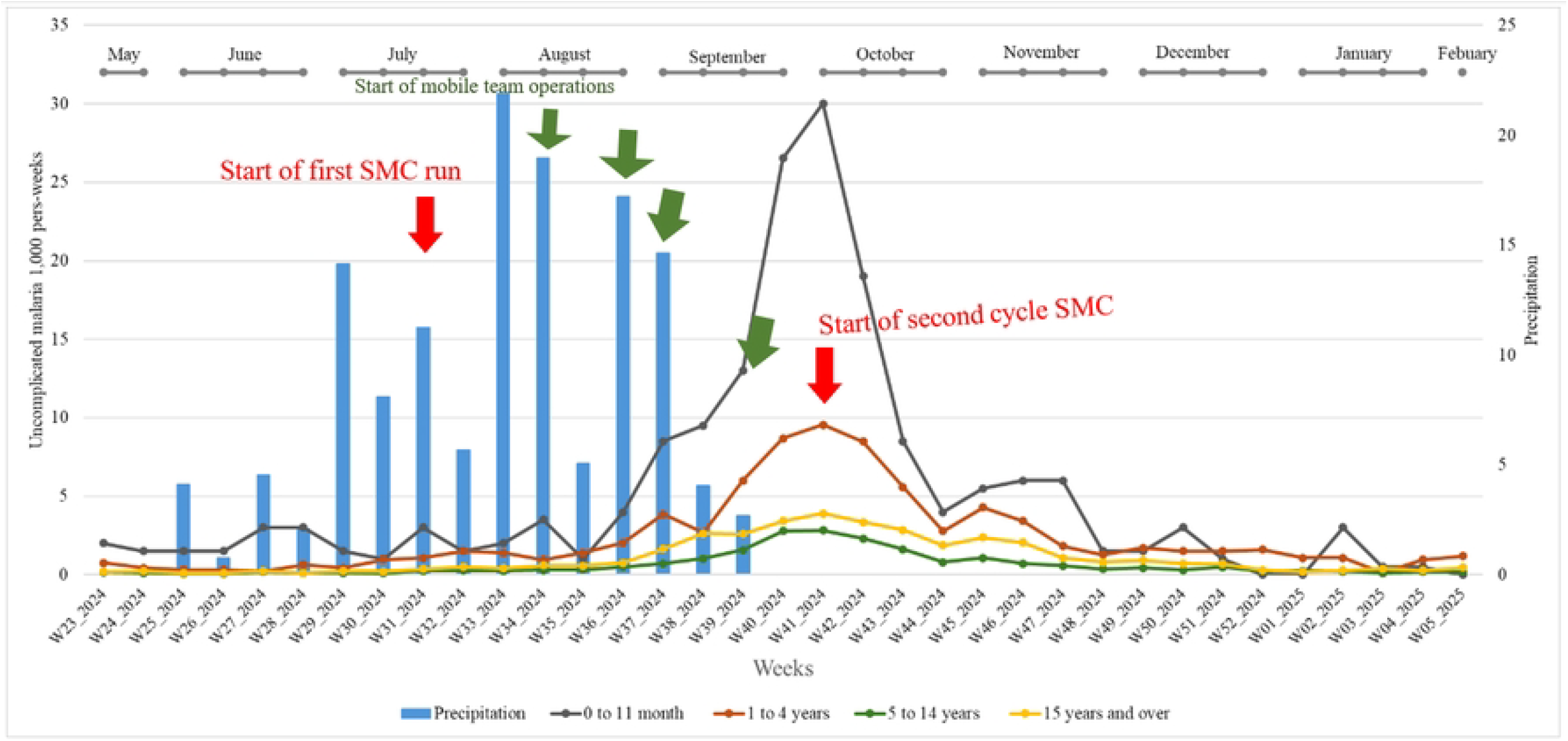
Trends in weekly incidence of uncomplicated malaria by age group and rainfall from S23 (May-2024) to S5 (February-2025).

The incidence of uncomplicated malaria in children aged [0-11] months per 1,000 person-weeks is represented by the black line, while that in children aged [1-4] years is shown in red. Incidence in the [5-14] age group is shown in green, and in the 15+ age group in dark yellow. Weekly cumulative precipitation (mm) is shown on the blue bar chart.

### 3.3. General description of the time series of severe malaria incidence by age group

Out of a total of 2,310 confirmed cases of severe malaria, the age groups most affected were children aged [0-11] months and [1-4] years, with incidence rates of 1.48 and 0.97 per 1,000 person-weeks respectively. On the other hand, the age groups least affected were those [5-14] years, with incidence rates of 0.32. It is worth noting that people aged 15 and over presented many severe cases, with incidence rates of 0.78 per 1000 person-weeks, and a high peak of 355 severe cases, in week 39 (3rd week of September). This 15+ age group showed a severity rate of 0.59 severe cases per 100 malaria cases, higher than the other severity rates of 0.04; 0.13; 0.22 for [0-11] months; [1-4] years and [5-14] years respectively.

**Figure 3:**
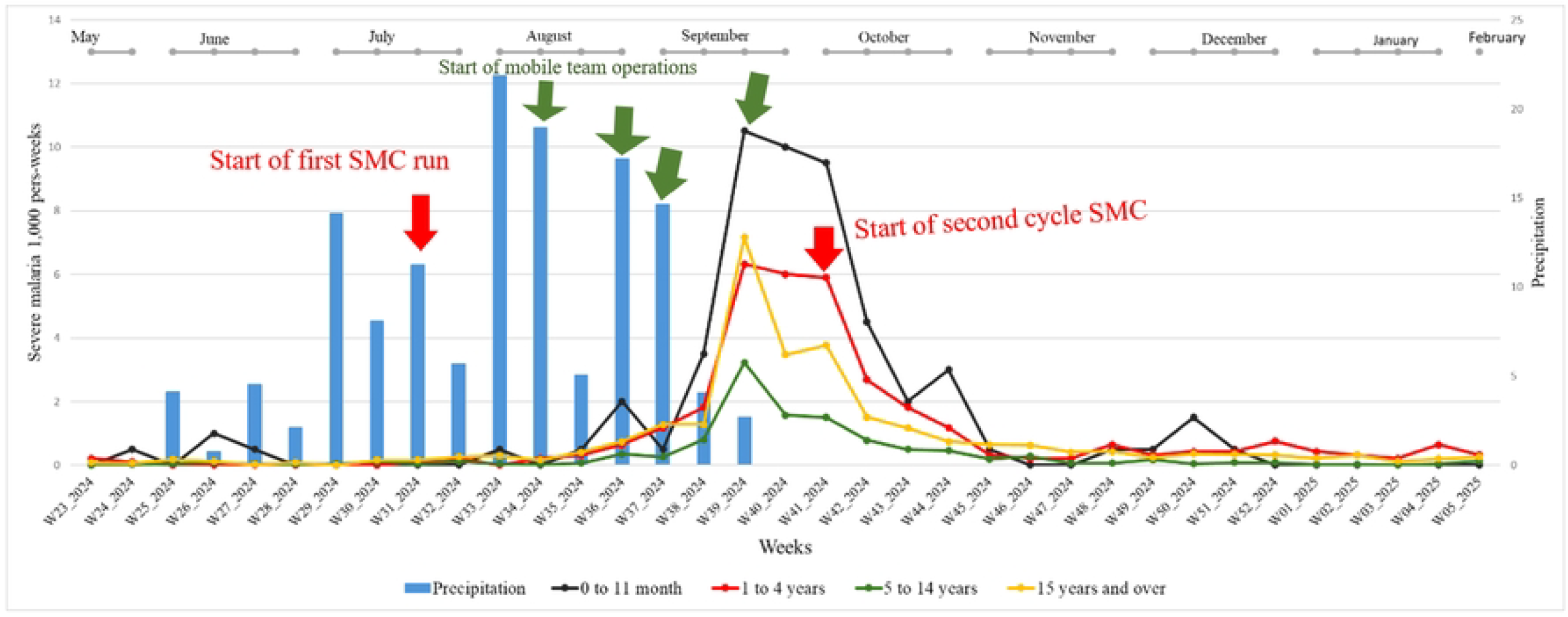
Trends in weekly incidence of severe malaria by age group and rainfall from S23 (May-2024) to S5 (February-2025).

Incidence per 1,000 person-weeks is differentiated by age group: children aged [0-11] months are represented by the black curve, those aged [1-4] years by the red curve, those aged [5-14] years by the green curve and those aged 15 years and over by the dark yellow curve. Weekly cumulative precipitation (mm) is represented by the blue bar chart.

### 3.4. Map of the estimated population at risk of malaria

Figure 4 shows the spatial distribution of the estimated population at risk. Due to a very small and scattered estimated population in the Tin-Essako health district and in the northwest and southwest of the Tessalit health district, the risk of malaria was low in these areas. The risk of malaria was particularly high in the north of the Abeibara district, on the border with Algeria, and in the southeast of the same district, where the estimated population was 9,449 people over 16,328 km^2^. In contrast, in areas close to the Kidal and Tessalit health districts, the risk of malaria was lower, with 2,066 people over 1,916 km^2^. The estimated population was low in the north of the Tessalit health district, on the border with Algeria, but higher in the south, on the border with the Kidal health districts and in the center of the Tessalit district, where the population was 17,624 over 21,535 km^2^. The Kidal health district was the most at risk for malaria cases, with populations scattered throughout the district. According to estimates, the highest population numbers were observed in the center of the Kidal health district, with 57,724 people over 13,323 km^2^.

**Figure 4:**
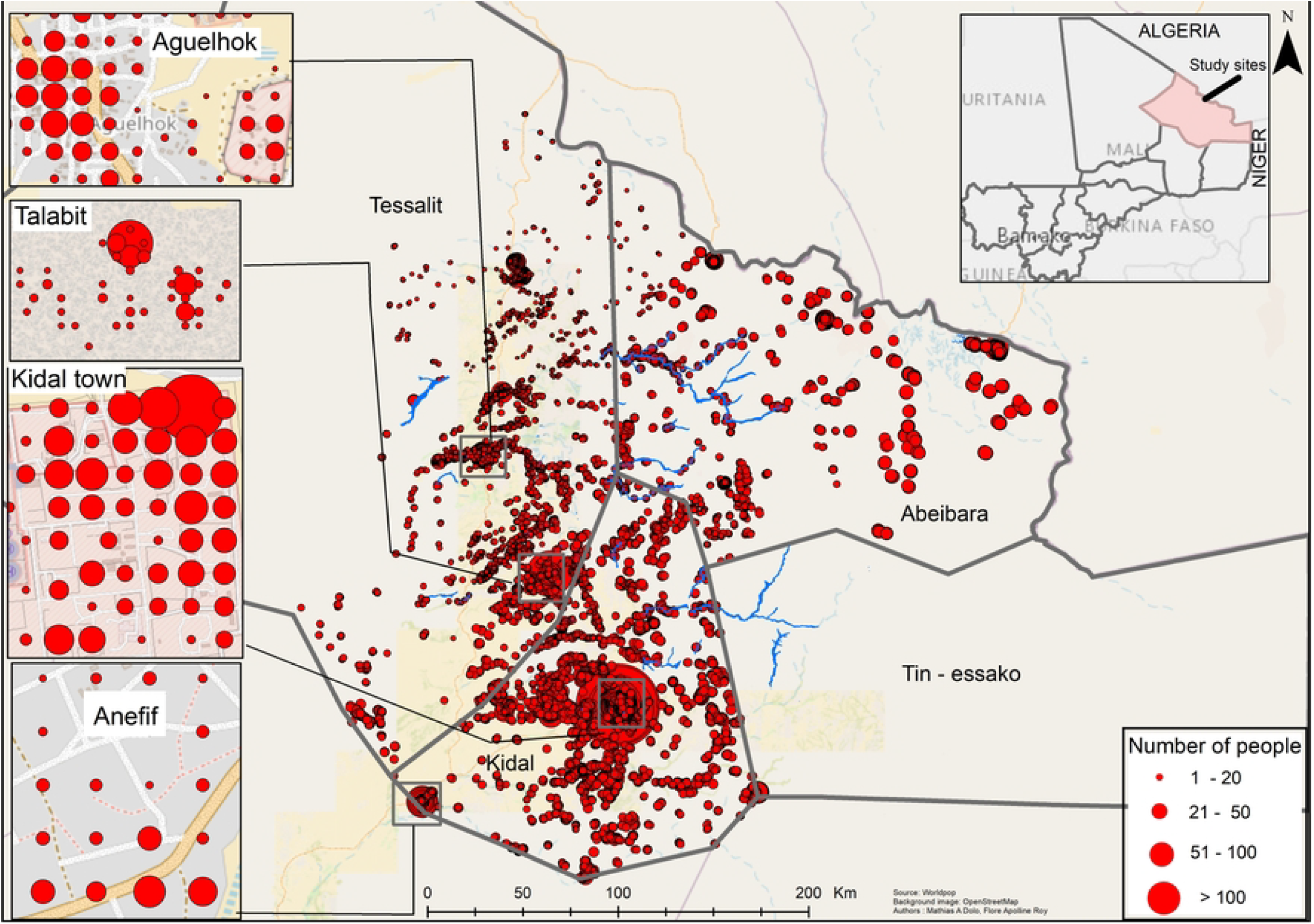
Map showing the distribution of estimated population numbers in districts where we have identified at least 1 case of malaria. The size of the red dots varies according to the estimated population, ranging from 1 to 154 people.

### 3.5. Response to the outbreak

Facing this malaria outbreak, the Kidal Regional Health Department, in collaboration with the National Malaria Control Program (Ministry of Health) and its partners, and the General Directorate of Health and Public Hygiene, undertook two types of intervention:

- Mobilization of 36 community health workers were mobilized for the 2nd seasonal malaria chemoprevention (SMC) campaign, aimed at children under 5 and extended to children up to 15. The first round, aimed at children under 5, took place before the epidemic, from July 29 to August 2, 2024 (31st week), treating 6,236 children. The second run took place from October 6 to 10, 2024, corresponding to the 41st week, and treated 27,334 children aged 0 months to 15 years.
- Mobilization of seven mobile teams, each comprising a driver, a doctor, a midwife, a nurse and a community mediator, were deployed to provide (free for all) diagnosis (RDT) and treatment (Artesunate inj 30 mg and 60 mg, Artemether inj 20 mg and 40 mg, quinine 400 mg and 200 mg and CTA Child, Adolescent and Adult, Sulfadoxine/Pyrimethamine, Paracetamol cp 500 mg and syrup) of malaria cases. 5 teams were deployed to remote areas in the Kidal district health units, and 2 teams were sent to Internally Displaced People in the Kidal district. Over the whole period, 1561 cases of malaria were treated by mobile teams.

#### 3.5.1. Regression modal

The full model showed a significant effect of the 2 interventions slowing down the malaria epidemic, with a SIR = 0.5 CI95[0.33; 0.75] (p=0.002) and 0.48 CI95[0.28; 0.82] for SMC and mobile teams. The R2 of the model was 99.7%, while the model without intervention showed an R2 of 91%. The Simulations carried out with the 2 models show that without intervention, the start of the malaria epidemic would have been exponential (median error −11 cases, range [−712; 12546], total number of predicted cases: 26071).

## 4. Discussion

In the Kidal region, a malaria epidemic followed flooding in the region, from week 23 of 2024 (3rd week of May) to week 5 of 2025 (1st week of February), the incidence rate of 1,688 /1000 person-week was well above the previous 3 years of 2021, 2022 and 2023 (resp. 0.412; 1.154; 0.706) according to a report by the national health information system. Cumulative rainfall was

135.11 mm over 15 weeks (June to September), with rain falling every week. The highest amount of rainfall was recorded to August, with 40.84 mm in 14 days (10) for previous years, with breaks in observation between weeks. It should be noted that the heavy rains came on top of several malaria cases already reported, a sign of low but already present circulation. Children aged 0-11 months and 1-4 years were most affected (14). However, we observed a higher severity rate in the over-15s, which is contrary to the malaria profile in endemic areas (15). Several studies support the idea that adults should be considered as a population to be closely monitored in epidemic malaria zones, due to their low exposure to malaria (16,17). This shows the importance of chemoprevention for adults at high risk of malaria. As a result, the burden of malaria and its evolution over time remain poorly understood in adults, and need to be studied in depth in areas with epidemic profiles.

It should be noted that the number of malaria cases is probably underestimated. Indeed, the interruption of the service for security reasons may have affected the reporting of the number of cases and the coverage of access to care. Despite these limitations, the health services were able to maintain care for population.

Due to the problems of insecurity in the Kidal region since 2013, as well as extreme rainfall events in August 2024, supply and health service circuits have been interrupted (7). However, the lack of regular malaria chemoprevention coverage could largely explain the high malaria incidence rate observed in children under 5 in our study.

The Kidal health district zone was the most at risk from this epidemic, as it has a larger population, in addition to the migration center for nomadic populations and the sites of the region’s 3 other health districts. This concentration of populations potentially aggravated the situation, making the management of the health crisis more complex. These results corroborate (19) that modifiable risk factors such as overcrowding were associated with increased malaria incidence in Nigeria.

Extreme and localized weather events can be the cause of serious malaria outbreaks. At present, the country’s northern regions have few mechanisms in place to ensure continuity of health services for the prevention and management of cases among vulnerable populations.

The mobilization of the regional health department (7), under the coordination of national, the second round of chemoprevention of seasonal malaria was organized (week 41) from October 6 to 10, 2024, and mobile teams were deployed in areas far from health facilities and in camps for internally displaced persons to provide free care from August 10 to September 20. The results of our model confirm that the interventions have significantly reduced malaria morbidity. These results corroborate (19,20,21) on the implementation of interventions to reduce malaria. Moreover, since malaria is already circulating at a low level before the arrival of the rainfall, it is possible to propose a mass treatment strategy as soon as the first rains arrive to break the booster effect, or, at least, to prepare diagnostic and treatment inventories.

Outside periods of crisis, as described in this work, current malaria control strategies in the Sahelian zone mainly involve the use of insecticide-treated mosquito nets, at intermittent preventive treatment for pregnant women and chemoprevention of seasonal malaria (SMC) in children under 5 years of age by community health workers when they are present. While the presence of community health workers needs to be strengthened, the security context remains an obstacle. In these contexts of both security and meteorological crisis. Mobile teams play a vital role in reducing malaria. Especially since the impact of the mobile teams’ interventions has been confirmed in several studies where the health system has reached its limits (22,23).

The fight against malaria is going through a turbulent period, with an accumulation of crises that threaten to reverse decades of progress (24). The Sahelian countries hard hit by the disease are facing serious budget deficits and unprecedented security and meteorological crises (24). According to the IPCC report, it is predicted that the countries of the Sahel will experience extreme rainfall events and flooding from both rain and rivers (3). Several studies (25,26) showed the significant increase in malaria after the floods. According to the scenarios (27) the duration of drought in West Africa is set to increase by around 4 months over the coming years. The increased intensity of extreme rainfall events combined with the increase in drought episodes will increase and modify malaria trends in the Sahel (28). Another study conducted in the 43 countries of the WHO African region (29) confirms that an increase in extreme rainfall events will increase the number of malaria cases in West Africa by 51.3 million by 2080.

## Conclusion

In short, extreme weather events, will have a significant impact on unstable malaria in the Sahelo-Saharan region. Events such as the floods mentioned here represent major challenges for national programs, requiring urgent responses from health authorities to put in place appropriate control strategies such as mobilizing mobile teams, extending community health wokers, extending SMC to other age groups, improving stock management at the end of the dry season, and strengthening case management (free diagnosis and treatment for all) in areas affected by epidemics.

## Financing

This work was funded by a French ARTS grant from the French Institute for Development Research France (IRD), and supported by the NGO Prospective & Cooperation. We would also like to thank the partners who funded the interventions, notably the World Health Organization (WHO), the United Nations Children’s Fund (UNICEF), Doctors Without Borders (MSF) Spain and DDRK (Sustainable Development in the Kidal Region).

## Author contributions

MS, AT and KK collected the primary data;, AMD, MC, IS and JG designed the study; MS, AT, conducted the field study coordinated by AT; AMD and MC developed the statistical analysis plan with the participation of IS carried out the statistical analysis under the supervision of JG; AMD carried out the cartographic analysis with the participation of FAR and IS; MC, JG, CATT, MS, AT, AK, MM and KK participated in the statistical analysis and validated the results; AMD, MC, IS and JG drafted the manuscript. All authors have read and approved the final manuscript.

## Data availability

I have attached a supplementary file with associated information used in the manuscript.

## Acknowledgements

The authors would like to thank all the staff of the National Malaria Control Programme in Mali, and all the staff of the Kidal Regional Health Directorate, for their contribution to epidemiological surveillance.

